# Predicting diagnostic conversion from major depressive disorder to bipolar disorder: an EHR based study from Colombia

**DOI:** 10.1101/2023.09.28.23296092

**Authors:** Susan K. Service, Juan De La Hoz, Ana M. Diaz-Zuluaga, Alejandro Arias, Aditya Pimplaskar, Chuc Luu, Laura Mena, Johanna Valencia, Mauricio Castaño Ramírez, Carrie E. Bearden, Chiara Sabbati, Victor I. Reus, Carlos López-Jaramillo, Nelson B. Freimer, Loes M. Olde Loohuis

## Abstract

Bipolar Disorder (BD) is a severe and chronic disorder characterized by recurrent episodes of depression, mania, and/or hypomania. Most BD patients initially present with depressive symptoms, resulting in a delayed diagnosis of BD and poor clinical outcomes. This study leverages electronic health record (EHR) data from the Clínica San Juan de Dios Manizales in Colombia to identify features predictive of the transition from Major Depressive Disorder (MDD) to BD.

Analyzing EHR data from 13,607 patients diagnosed with MDD over 15 years, we identified 1,610 cases of conversion to BD. Using a multivariate Cox regression model, we identified severity of the initial MDD episode, the presence of psychosis and hospitalization at first episode, family history of mood or psychotic disorders, female gender to be predictive of the conversion to BD. Additionally, we observed associations with medication classes (prescriptions of mood stabilizers, antipsychotics, and antidepressants) and clinical features (delusions, suicide attempt, suicidal ideation, use of marijuana and alcohol use/abuse) derived from natural language processing (NLP) of clinical notes. Together, these risk factors predicted BD conversion within five years of the initial MDD diagnosis, with a recall of 72% and a precision of 38%.

Our study confirms many previously identified risk factors identified through registry-based studies (such as female gender and psychotic depression at the index MDD episode), and identifies novel ones (specifically, suicidal ideation and suicide attempt extracted from clinical notes). These results simultaneously demonstrate the validity of using EHR data for predicting BD conversion as well as underscore its potential for the identification of novel risk factors and improving early diagnosis.

## Introduction

Bipolar Disorder (BD) is a common, highly heritable, chronic disorder characterized by episodes of depression and mania (or hypomania) (McIntyre et al., 2020). The diagnosis of BD is challenging in clinical practice, with a mean delay between illness onset and diagnosis of 7 years (Scott et al., 2022). The main reason it takes such a long time to reach a BD diagnosis is that illness onset is often marked by a depressive episode (Angst & Sellaro, 2000; Daban et al., 2006; Kalman et al., 2021), resulting in a diagnosis of unipolar major depression (MDD) in 60% of BD patients. The delayed diagnosis of these BD patients has many potentially detrimental consequences, including prescription of antidepressants in the absence of mood-stabilizing drugs, which in some cases can lead to mania, poor clinical outcomes and, consequently, high health care costs (McIntyre et al., 2020). Reducing the time to a BD diagnosis would thus be of great benefit to patients, their families, and society.

Several studies have tried to determine factors that are predictive of conversion from MDD to BD. The meta-analysis of Kessing et al., (Kessing et al., 2017) examined 31 different studies and could not identify risk factors that acted consistently across studies; they attributed their lack of findings to differences among studies in methodology. In another meta-analysis Ratheesh et al., (Ratheesh et al., 2017) examined 56 studies (19 overlapping with (Kessing et al., 2017)) and found family history of BD, an earlier age of onset of depression, and presence of psychotic symptoms all to be significant predictors of conversion to BD. However, most existing studies rely on small cohorts, include few predictors, and/or rely on patient recall.

Two analyses of registry data from Denmark (Musliner & Ostergaard, 2018) and Finland (Baryshnikov et al., 2020) are notable for their large samples of consistently ascertained and evaluated individuals. Based on analyses of registry data from 91,587 Danish residents with a diagnosis of MDD, Musliner & Ostergaard (Musliner & Ostergaard, 2018) found family history of BD, psychotic depression, prior diagnoses of non-affective psychosis, inpatient or emergency room treatment at the first MDD episode, previous diagnosis of alcohol abuse, female sex, and depression severity, all to be significant predictors of conversion to BD. In a sample of 43,495 Finnish residents hospitalized with unipolar depression, Baryshnikov et al., (Baryshnikov et al., 2020) also found female sex, the type and severity of first MDD episode, and the age of the first MDD episode to all be predictive of BD conversion.

National registry data, such those used in Musliner & Ostergaard (Musliner & Ostergaard, 2018) and Baryshnikov et al., (Baryshnikov et al., 2020), are extremely valuable because they represent complete information from an entire country, uniformly recorded and longitudinal in nature. While national registries are only available in a select few upper income countries, electronic health records (EHR) have become widely available in recent decades, including in lower-middle income countries (LMIC). EHR data share characteristics of registry data: they are systematic records of health care utilization and longitudinal in nature. In addition, in settings where all members of a population have equal access to health care, and catchment areas are well-defined, EHR data may approach registry data in terms of their potential for large epidemiologic investigations that include a time component. Moreover, the breadth and granularity of EHR data provides information beyond data available in registries: e.g., EHR include clinical notes describing the progression of clinical features recorded at every visit, and daily during a hospital stay. Features extracted from these notes provide additional layers of information above the structured data types usually available in registries.

The Clínica San Juan de Dios Manizales (CSJDM) in Manizales, Colombia, is the primary psychiatric hospital for the entire department (state) of Caldas (population 1 million). EHR data have been available since 2005, and treatment is available to all residents regardless of insurance status (Song et al., 2022). We previously validated information related to diagnoses in the records and established a Natural Language Processing (NLP) pipeline for the reliable and precise extraction of symptoms and behaviors from the clinical notes (Hoz et al., 2022). We further showed how the longitudinal EHR data including diagnoses and clinical features can be used to delineate psychiatric patient trajectories and to study factors associated with diagnostic stability.

Here, we aim to identify factors associated with the diagnostic switch from MDD to BD using a multivariate Cox regression model based on features extracted from the CSJDM EHR. Similar to Musliner & Ostergaard (Musliner & Ostergaard, 2018) and Baryshnikov et al., (Baryshnikov et al., 2020), we find that severity of the first MDD episode, presence of psychotic symptoms recorded in ICD codes, family history of mood or psychotic disorders, history of psychiatric treatment as a minor, and female gender all to be predictive of conversion from MDD to BD, thus highlighting the validity of using EHR. In addition, we also observe associations with medication classes and NLP-derived clinical features not available in registry data: anti-psychotic and mood-stabilizing medication use, as well as delusions, use of alcohol, suicide attempt and suicidal ideation are all risk factors for conversion to BD, while anti-depressant and marijuana use are protective factors. We further test the ability of our model to predict which patients newly diagnosed with MDD will convert to BD within 5 years.

## Methods

### Sample

Participants in this study were identified through electronic health records (EHR) at the CSJDM in Manizales, the capital of the department (state) of Caldas. The CSJDM is the primary mental health care facility in the department (state) of Caldas, Colombia; it serves all inhabitants of Caldas, regardless of health insurance status. CSJDM has maintained EHR on all inpatient and outpatient visits since 2005. We used EHR information entered in the system from inception in 2005 through December 31, 2021, in our analyses.

We extracted from the EHR information on patients, who at some point in their time course, had an inpatient or outpatient International Statistical Classification of Diseases and Related Health Problems, 10th Revision (ICD-10) code indicating a diagnosis of major depressive disorder (MDD: F32 or F33). If patients received both an MDD and a bipolar (BD: F31) diagnoses through their time course, we included only patients whose MDD diagnoses preceded their BD diagnosis. Patients that transitioned from MDD to BD were included if they did not transition back to an MDD diagnosis later in their time course. Patients with an ICD-10 diagnoses of schizophrenia (SCZ) or schizoaffective disorder (at any point in their time course) were excluded, as were patients whose first MDD diagnosis was before age eight. To be included in the analysis, patients had at least one day of follow-up after their initial MDD diagnosis.

Ethical approval for this study was granted by The Institutional Review Board, Medical Institutional Review Board 3, at UCLA; the Comité de Ética del Instituto de Investigaciones Médicas, at Universidad de Antioquia; and the Comité de Bioética de Clinica San Juan de Dios, at CSJDM.

### Data Extraction

The EHR data at CSJDM are composed of both structured and unstructured information and are contained in two different databases: the first one spanning the years 2005-2015 and the second one 2016-present day. The structured fields include demographics such as age and sex, vitals, medications, diagnostic codes (ICD-10), health system utilization data such as the duration and type of encounters (inpatient, outpatient, emergency room), among others. From the structured data, any field considered to be Protected Health Information by HIPAA (Office for Civil Rights, 2023) was removed from the records. In addition, names and numbers exceeding 5-digits (potential ID numbers) were stripped from the text using regular expressions.

The unstructured part of the EHR is composed of diverse types of notes, as described in De la Hoz et al., (Hoz et al., 2022). We extracted information on symptoms, behaviors, substance use, and family history of psychiatric disorders from the unstructured part of the EHR using Named Entity Recognition (NER) and Negation Detection (Hoz et al., 2022), In comparison to manual chart review of 105 patients by psychiatrists at CSJDM (our gold standard), our data extraction methods for unstructured data have recall ranging between 0.67 and 0.86 and precision ranging between 0.75 and 0.86 (Hoz et al., 2022).

### Predictors

Predictors were based on data extracted on or before the first visit to the CJSDM with an MDD diagnosis, and can be grouped into broad categories of demographic, family history of psychiatric disorders, severity of the first MDD diagnosis, psychiatric diagnostic history, substance use, prescription medication use, and symptoms/behaviors. Demographic variables included the age at the first MDD diagnosis, sex, and residence. Residence was coded using two dummy variables: residence in Manizales or residence in the outlying municipality of Aranzazu, a community 55 km from Manizales shown to have an extraordinarily high incidence of BD (Song et al., 2022), We coded information on family history of psychiatric disorders into four dummy variables: history of BD, SCZ, MDD, or Psychosis. The severity of the first MDD episode was captured by the type of first MDD episode, and coded into two indicator dummy variables: hospitalized at the first MDD diagnosis and seen in the emergency room at the first MDD diagnosis (without subsequently being hospitalized). The type of first MDD episode (as indicated by the three-digit ICD-10 code) was categorized into two dummy variables: Severe No Psychosis, Severe with Psychosis, with all other MDD diagnoses as reference. Data on patient psychiatric history was coded by two dummy variables, any psychiatric diagnosis before the first MDD diagnosis, and an indicator if the first psychiatric visit of any type was while the patient was a minor. Substance use was coded as five dummy variables that recorded any use (not necessarily current use) of tobacco, alcohol, marijuana, cocaine, or other recreational drugs, up to and including the time of the first MDD diagnosis. We condensed use of prescription medications into five dummy variables: use of any anti-depressant, use of any anti-psychotic, use of any mood stabilizer, use of any hypnotic/anti-anxiety, and use of any hypothyroid medication, up to and including the time of the first MDD diagnosis. We focused on three symptoms/behaviors that were reliably extracted in De la Hoz et al., (Hoz et al., 2022): suicide attempt, suicidal ideation, and presence of delusions. Each was coded as a dummy variable, and recorded presence/absence of these symptoms/behaviors at any point up to and including the time of the first MDD diagnosis.

### Outcome Measures

For each patient we recorded the time, in days, from their first MDD diagnosis to their conversion to a BD diagnosis. If the patient did not convert to BD before December 31, 2021, they were considered to be censored for the outcome, and we record the time, in days, from their first MDD diagnosis to the last known visit in the EHR.

### Statistical Analysis

Analyses were conducted in R version 3.6.1. We divided our data into a training set (70%) and a testing set (30%), preserving the ratio of censored observations to BD conversions in the division. We performed analyses in the training data and evaluated model predictions in the test data. We used multivariate Cox regression (implemented in the survival package, (Therneau, 2022)) to estimate the hazard ratio for each predictor. To evaluate the Cox regression assumption of proportional hazards we performed a generalized linear regression of the scaled Schoenfeld residuals on functions of time, using the cox.zph() function in the survival package. A non-zero slope is an indication of a violation of the proportional hazard assumption.

In the held-out test data we used the hazard ratios estimated from the multivariate model (developed in the training data) to calculate the probability of converting to BD within five years (hereafter abbreviated as PrC5) of the first MDD diagnosis for each patient. We chose five years as our primary follow-up time, as we observed 85% of observed conversions to occur within this time frame and a five-year follow-up is a common time frame in survival analysis. We also evaluated model performance one and two years after the first MDD diagnosis, as these were the times of median and mean, respectively, observed conversions (see Results).

We can apply a decision threshold to the PrC5 to assign a label (converter/non-converter) to the patients in the held-out test data. The chosen threshold determines the balance between the number of false positives and false negatives resulting from our classification. The area under the ROC curve (AUC) is a measure of the performance of these conversion probabilities to classify new observations. The AUC is equal to the probability that a randomly chosen BD converter will have a higher PrC5 than will a randomly chosen non-converter.

Key to evaluating performance of the PrC5 from our Cox model in correctly classifying new observations is knowing the true status of our patients (converter or non-converter). We cannot know this with certainty for censored observations, we only know that the patient had not converted at the last observation time. At each time point *t*, it would be tempting to discard patients who were censored before time *t* and classify the remaining patients as converted, if they transitioned to BD on or before time *t*, non-converters if they had not converted by time *t*, and evaluate the recall and FPR on this subset of patients. This naïve approach has been shown to produce biased estimates of sensitivity and AUC, when censoring depends on the conversion probability (Blanche et al., 2013). We instead use the PrC5 in a nearest-neighbor weighted Kaplan Meier approach (Heagerty et al., 2000) to estimate the ROC curve, as implemented in the package survivalROC (Heagerty PJ, 2022). We evaluated the variability in estimates of recall, precision, and AUC using 1,000 different splits of our data into training/test data sets.

### Post-hoc analyses

While our primary model used data on predictor variables collected on or before the first MDD episode (termed our baseline model), we performed a secondary analysis using covariate data collected during interim visits to the clinic, after the first MDD diagnosis but before conversion to BD (or censoring). This secondary analysis employed time-dependent covariates in a Cox model. We compared hazard ratios estimated in the training data in the time-dependent analysis to those estimated in the baseline model to evaluate the possible gain in predictive power by using data collected on interim visits.

To assess additional predictive power using non-linear models we performed an additional secondary analysis using random survival forests as implemented in the R package ranger (Wright, 2017). A forest of 300 trees was grown using the training data, and model predictions evaluated in the held-out test data.

## Results

### Sample Characteristics

The EHR contained records on 73,785 patients seen between 2005 and December 31, 2021. After applying inclusion and exclusion criteria described in the Methods, our final sample for analysis was comprised of 13,607 patients (Supplementary Figure 1). The sample was followed for a total of 21,573.8 person-years; length of follow-up ranged from 1 day to 15 years (mean: 1.6 years; SD: 2.4 years). A total of 1,610 patients converted to BD during follow-up; on average they convert within 2.1 years of their MDD diagnosis (SD=2.7 years) and 49% convert within one year of diagnosis (Figure 1A). The Kaplan-Meier estimate of the survival curve indicates the highest incidence of conversion to BD within the first year of the MDD diagnosis (Supplementary Figure 2).

**Figure 1.**
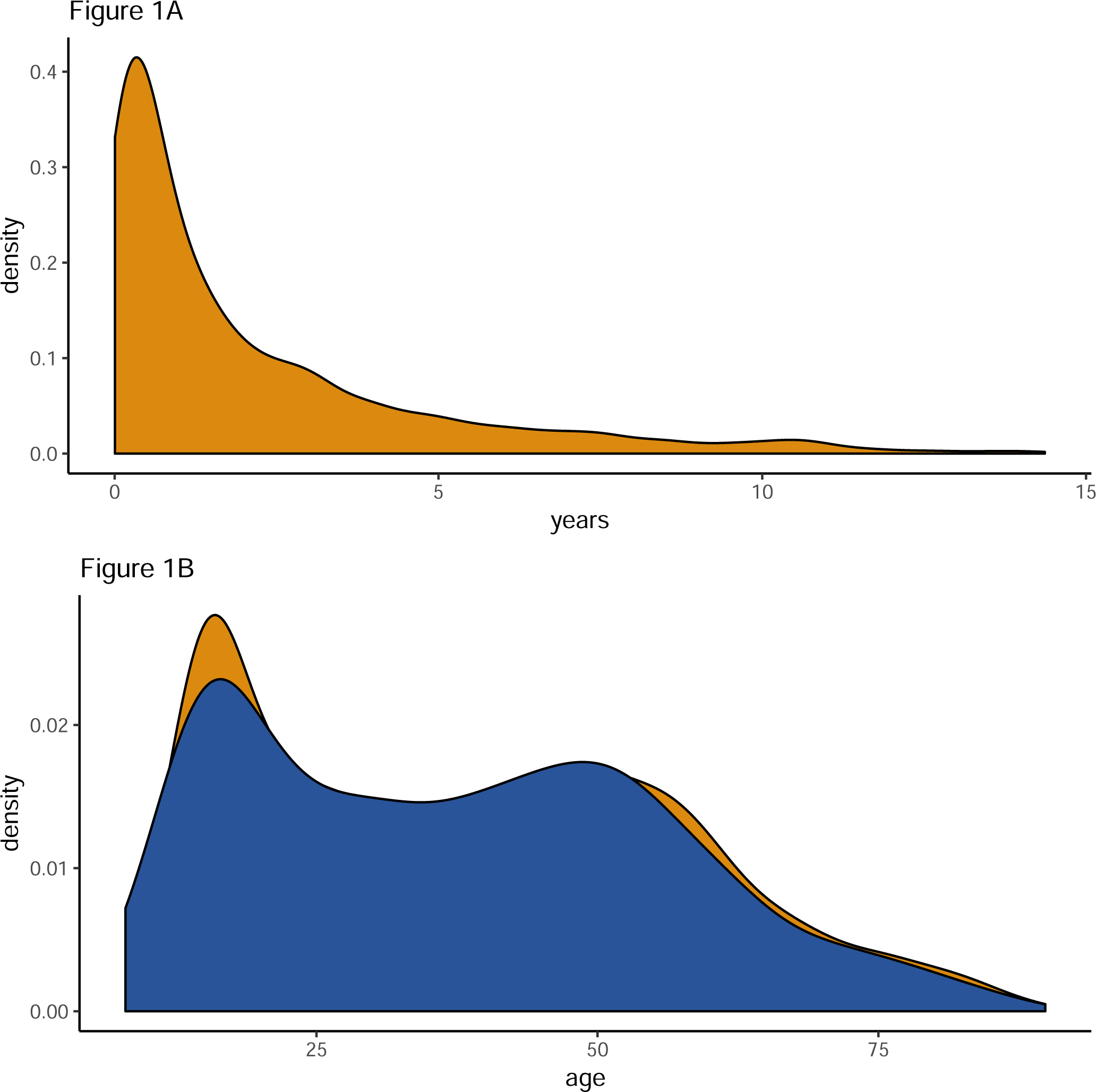
(A) Distribution of time to convert to BD after the initial MDD diagnosis for 1,610 converters. (B) Distribution of age at first MDD for 1,610 converters (blue) and 11,997 censored patients (green).

The sample was majority female (66%), primarily from Manizales (68%), with relatively low rates of substance use. Most patients (72%) were on anti-depressants on or before their first MDD visit. The distribution of the age of patients at their first MDD diagnosis is bimodal (Figure 1B), with peaks in the late teens and at mid-life. A full description of the prevalence and distribution of predictor variables for the sample can be found in Table 1.

**Table 1.**
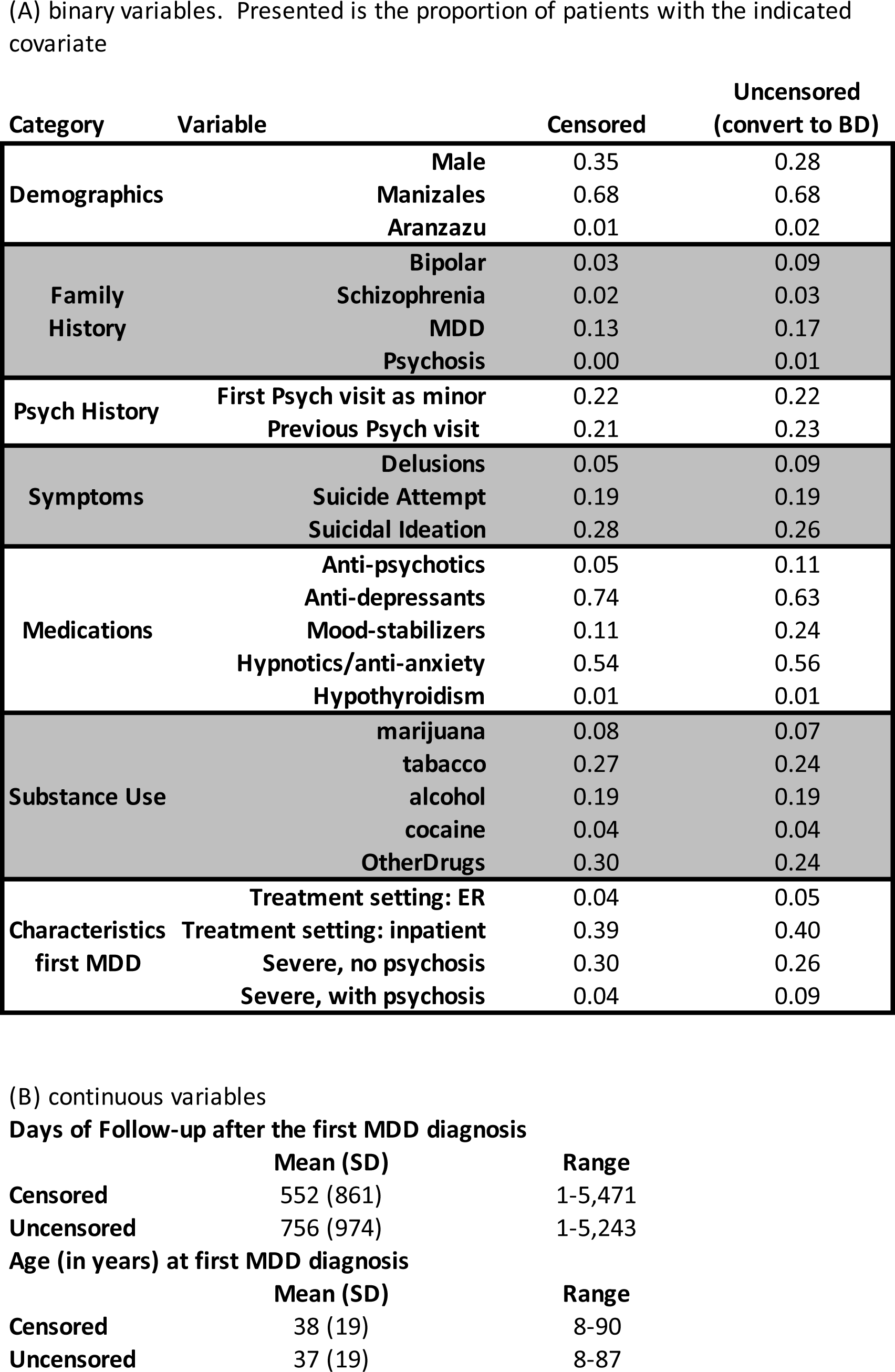
Sample characteristics of censored (n=11,997) and uncensored (n=1,610) observations in the full sample.

### Cox Model on Training Data

We tested the assumption of proportional hazards in the training data for each of the 27 predictor variables. Correcting for multiple testing, we found one variable (use of mood stabilizing drugs) to be significant at the 0.05 level. Inspection of the residual plot, however, showed the deviation to be very modest (Supplementary Figure 3).

The baseline Cox regression model used 9,525 patients in the training data: 1,127 converted to BD and 8,398 were censored (Figure 2, Supplementary Table 1). Males had a reduced rate of conversion to BD compared to females (hazard ratio [HR]: 0.69; 95% CI 0.61-0.80). Participants from Manizales (the city where the CSJDM is located) had a reduced rate of conversion to BD compared to participants from outside of the city (HR: 0.83, 95% CI 0.73-0.94). Delusions, suicidal ideation, or a previous suicide attempt were associated with increased the rate of conversion to BD a similar amount (delusions HR 1.26, 95% CI 1.01-1.58; ideation HR 1.27, 95% CI 1.09-1.47; attempt HR 1.22, 95% CI 1.031-1.44).

**Figure 2.**
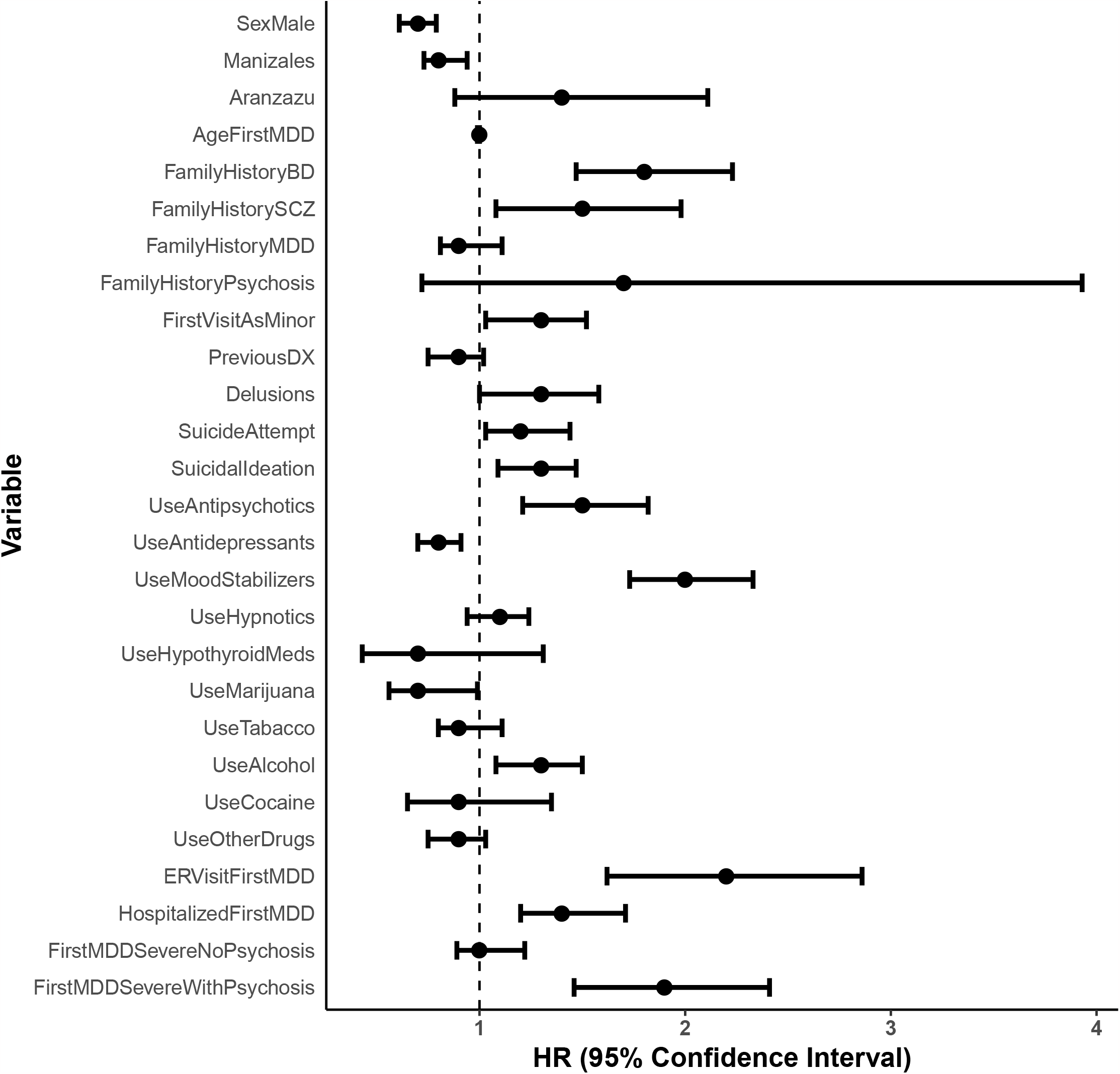
Hazard ratios and 95% confidence intervals from the baseline multivariate Cox model used on the training data.

Use of anti-psychotics or mood stabilizers on or before the first MDD diagnosis was associated with increased rate of conversion to BD (anti-psychotic HR: 1.48, 95% CI 1.21-1.82; mood stabilizer HR: 2.01, 95% CI 1.73-2.33), while use of anti-depressants with a decreased the rate of conversion (HR: 0.80, 95% CI 0.70-0.91). Of the substance use variables, alcohol was associated with an increased rate of BD conversion (HR: 1.27, 95% CI 1.08-1.50) and marijuana use was associated with a decreased rate of BD conversion (HR: 0.74, 95% CI 0.56-0.99). While the age at the first MDD diagnosis was not associated with conversion to BD, having a visit to the psychiatric hospital as a minor (not necessarily for MDD) was associated with increased rate of conversion to BD (HR: 1.25, 95% CI 1.03-1.52). Family history of BD or SCZ increased the rate of conversion to BD (BD HR: 1.81, 95% CI 1.47-2.23; SCZ HR 1.46 95% CI 1.08-1.98), while having a family history of MDD or psychosis was not strongly associated to BD conversion rate (Figure 2). Patients who visited the ER at their first MDD diagnosis, were hospitalized at the time of their first MDD diagnosis, or received a diagnosis of Severe MDD with psychosis, had increased BD conversion risk (ER visit HR: 2.15, 95% CI 1.62-2.86; Hospitalized HR: 1.43, 95% CI 1.20-1.70; Severe MDD with psychosis HR: 1.88, 95% CI 1.46-2.41).

Our finding that the of use of antidepressants is associated with a decrease in the rate of conversion to BD appears in contrast with previous work, where anti-depressant use in the absence of a mood stabilizer has been shown to induce manic episodes (Viktorin et al., 2014). In our sample, participants prescribed anti-depressants at the time of their first MDD episode are also less likely to have been hospitalized at that first episode, less likely to have had a severe MDD diagnosis (psychotic and/or non-psychotic), less likely to have had a suicide attempt, and less likely to experience delusions (summarized in Supplementary Table 2) than are participants that were not prescribed antidepressants; they are more likely, however, to have a family history of MDD. The apparent protective effect of anti-depressants in our sample may be due to milder presentation and lower prevalence of other risk factors in this group.

**Table 2.**
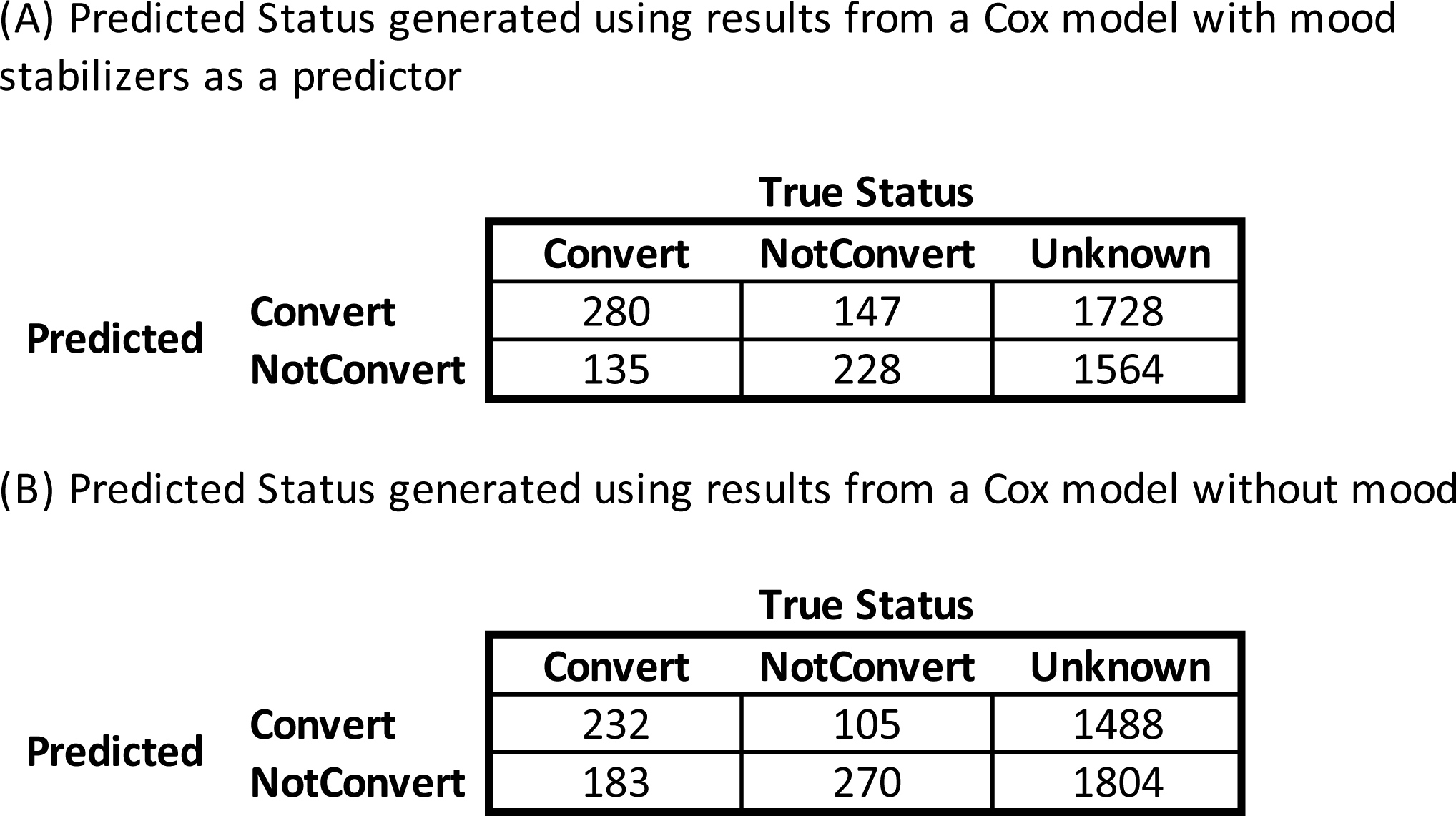
Predicted Status 5 years after the first MDD episode vs. True Status in 4,082 patients held out in a test data set.

### Prediction in held-out data

Next, we test how well our multivariate model can predict conversion to BD. We applied the results of the baseline Cox model developed in the training data to 4,082 patients (483 converters, 3,599 censored) held-out as a test data set, and estimated the probability of converting to BD five years after their initial MDD diagnosis (PrC5). We find the PrC5 to be higher in patients where we observed conversion to BD than in censored patients (Figure 3A). The cases in the top 10% of the PrC5 have ∼2x the number of observed converters than did the cases in the bottom 10% (83 vs. 39, respectively). We binned the PrC5 estimated in the test data into quartiles, and plotted Kaplan Meier survival curves for each quartile (Figure 3B). Indeed, participants with higher PrC5 were observed to have a higher rate of conversion to BD than those with a lower PrC5. Moreover, among the 483 converters in the test data, the median time to convert decreases with increasing PrC5 (Supplementary Figure 4). The area under the ROC curve was 0.65 (95% CI 0.62-0.68) and the area under the precision-recall curve was 0.39 (95% CI 0.34-0.42) (Supplementary Figure 5). That is, the probability that a converter has a higher probability of converting based on our model is 65%.

**Figure 3.**
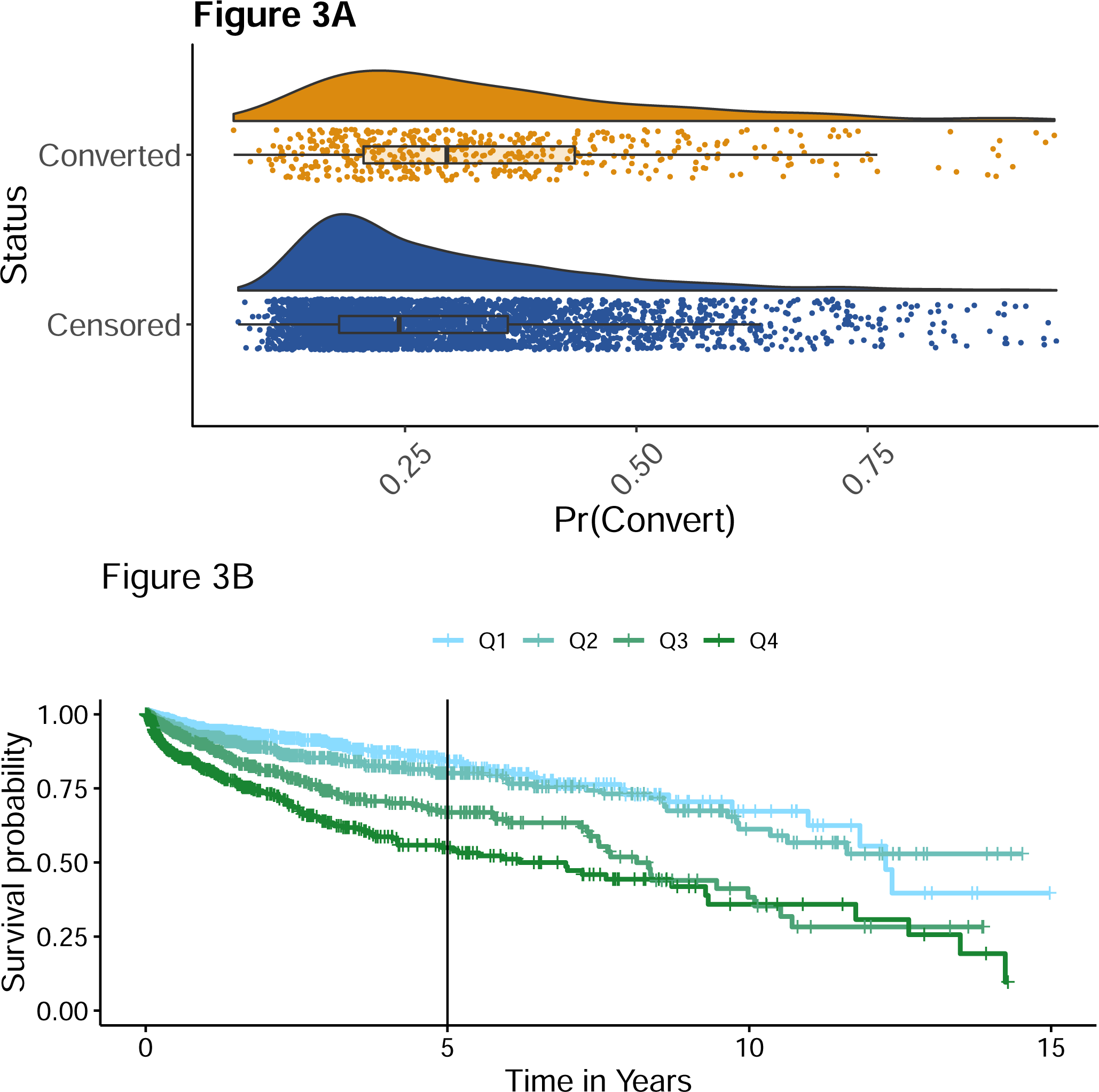
We estimated the probability to convert to BD within five years of the initial MDD diagnosis in 4,082 patients in the test data, using hazard ratios estimated in the training data. (A) Distribution of the probability to convert to BD in 483 converters and 3,599 censored patients in the test data. (B) Kaplan-Meier survival plots for 4,082 patients in the test data. Patients were split into quartiles based on the distribution of their probability to convert to BD within five years, using hazard ratios estimated in the training data. Q1=first quartile, Q2=second quartile, Q3=third quartile, Q4=fourth quartile.

At the maximum distance from the diagonal (the optimal probability threshold for classification) in Supplementary Figure 5, recall of conversion to BD within five years of the first MDD diagnosis was 0.72 (95% CI 0.60-0.84) and precision was 0.38 (95% CI 0.33-0.43). This recall was obtained at a PrC5 cut-point (to declare a patient to be a “converter” to BD) of 24%. At this probability threshold, we would capture 72% of patients that truly convert to BD within five years. This would come at the expense of modest precision: 38% of patients labeled as converters are observed to convert.

Using these PrC5 cut-points, we can label our 4,082 patients in the test data as predicted converters/non-converters, and compare to their observed status at each time point: converted to BD, not converted to BD, and unknown/censored (Table 2A). While these tables provide a useful visual of model performance in the test data, note that calculating recall and FPR from this confusion matrix by discarding the unknown/censored observations would result in a biased estimate of these parameters. In contrast, the recall estimates we report that were obtained from the weighted nearest-neighbor Kaplan Meier approach correctly handle the censored data.

Evaluating the ability of the model to identify converters after one or two, rather than five years results in a similar AUC (0.62 at one year, 0.64 at two years, 0.65 at five years), and recall (0.73 at one year, 0.76 at two years, compared to 0.72 at five years) and a substantially decreased precision (0.13 at one year, 0.21 at two years, compared to 0.38 at five years).

### Outliers in PrC5 in test data

As shown above, a higher probability of conversions was associated with an increase in the observed conversion rate in test data. However, there are patients who, despite having a high probability to convert (PrC5>50%, about twice the level we use to classify someone as a converter), did not convert. We hypothesized that they may still convert but do not have sufficient follow-up time in the EHR (Supplementary Figure 4). Indeed, among the 3,599 patients in the held-out test data that did not convert, 345 have PrC5>50%: for these patients, the mean follow-up time is 202 days shorter than those with PrC5<50% (mean 566 days (SD=867 days) vs mean 365 days (SD=735 days)). More generally, for every 10% increase in PrC5, we observe 87 days shorter follow-up time in non-converters (SE=9.5 days, p<2e-16), thus indicating that that while patients did not convert during the follow-up time, they may still convert in the future, possibly contributing to a modest AUC.

### The role of use of mood stabilizers in prediction

Our finding that prescription of mood stabilizers is a strong predictor of the rate of conversion to BD is not surprising, as this medication class is commonly used in patients with BD, and in the test data, participants using mood stabilizer have a higher PrC5 (Supplementary Figure 6). Given that some patients were prescribed mood stabilizers before their conversion to BD may indicate that the clinician suspected BD. In the full data with n=1,610 conversions, those on mood stabilizers convert an average of 208 days (0.6 years) earlier than do those not on mood stabilizers (mean 2.3 years (SD=2.7 years) vs mean 1.7 years (SD=2.4 years)). We ran a secondary analysis, omitting from the training data 1,199 patients (282 converters and 917 censored patients) that were on mood stabilizers on or before the time of their first MDD diagnosis. The Cox model assumption of proportional hazards was met in this secondary analysis. We find that for most coefficients, the magnitude of the HR in the two analyses is very similar (Supplementary Figure 7), however the smaller sample size reduced significance.

Using the HR estimated from a Cox model without mood stabilizer use as a predictor (but including the 1,199 patients on mood stabilizers), we evaluated the PrC5 in the held-out test data. The 4,082 patients in the held-out test data included 509 participants that were on mood stabilizers at the first visit (98 converters, 411 censored). We found that recall and precision estimated from the entire test data set were very similar to what we saw when we included mood stabilizer use in the model: recall=0.68; precision=0.39.

While it is not a formal clinician’s predictor for conversion to BD, we can treat a prescription of mood stabilizers prior to conversion as a proxy for such a predictor and compare it to the performance of our model that does not include the use mood stabilizers. We observe that the overall performance of this univariate prediction model is worse than our model excluding mood stabilizers (AUC is 0.54, precision is 0.38 and recall is 0.18).

### Value of data from interim clinic visits

HRs estimated from the Cox model using time-dependent covariates were similar in magnitude to those estimated using only predictor data gathered on or before the first MDD diagnosis in the baseline model (Supplementary Figure 8). A notable exception is the HR estimated for use of mood stabilizers, where the HR has increased to 3.8 (95% CI 3.32-4.39) and the confidence intervals from the two models did not overlap: patients who begin taking mood stabilizers after their first MDD diagnosis have a greatly increased risk of conversion to BD.

### Temporally independent prediction

We omitted from analyses patients who had no follow-up time after their first MDD diagnosis (Supplementary Figure 1). After our analyses of EHR data up to December 31, 2021 were complete, we evaluated whether any of these excluded patients had visited the clinic between Jan 1, 2022 and May 25, 2022 as a temporally independent prediction. We found that 50 patients had come back to the clinic in 2022, and four of these had converted to BD. Based on the data collected from their first MDD diagnosis and our baseline Cox model HR results, we estimated the PrC5 in these 50 patients, and found that the converters did have a somewhat higher PrC5 than the censored patients (Supplementary Figure 9), but the sample sizes are very small. Based on the 5-year prediction window and model settings described above, we correctly predicted conversion status for all three observed converters that converted within five years; one was observed to convert more than five years after their initial MDD diagnosis; that individual was predicted to be a non-converter.

Having shown before that distance to the hospital has an effect of treatment-seeking behavior for outpatients, specifically those with MDD (Song et al., 2022), we evaluated a model including only patients residing in Manizales, where the hospital is located. This model resulted in similar effect size estimates as well as predictive power (data not shown).

### Post-hoc analyses using random survival forests

To assess additional predictive power using non-linear models we performed a post-hoc analyses using random survival forests. Using this model did not improve performance compared to our Cox model (AUC=0.61, recall=0.38, precision=0.47; details not shown). In the random forest model, the most important variable to predict the outcome was the use of mood stabilizing drugs; the other top five variables include hospitalization at the first MDD episode, initial diagnosis of severe MDD with psychosis, a family history of bipolar disorder, and use of antidepressant drugs, all variables identified to be significantly related to conversion in the Cox model.

## Discussion

We show that by using data that could be collected at the time of the first MDD episode, we can recall 72% of patients that go on to convert to BD within five years. Our study, which relies entirely on EHR data from a psychiatric hospital, confirms many previously identified risk factors identified through registry-based studies (such as female gender and psychotic depression at the index MDD episode), and also identifies novel ones (specifically, suicidal ideation and suicide attempt extracted from clinical notes). We study the effect of mood stabilizers on our predictive models and quantify how risk factors identified *after* the index MDD visit but *before* conversion to BD differentially affect risk of converting to BD.

As in other studies (Baryshnikov et al., 2020; Kessing et al., 2017; Musliner & Ostergaard, 2018; Ratheesh et al., 2017; Scott et al., 2022), we found the highest incidence of conversion to BD within the first year of the MDD diagnosis; however, the conversion rates we observe are higher. Conversion rates in the first year after MDD diagnosis have been estimated to be ∼1.5% (Musliner & Ostergaard, 2018) to ∼4% (Kessing et al., 2017) our rate is 9.6% in the first year. We hypothesize that the rates are elevated in our sample because patients with mild MDD, who are less likely to convert to BD, may not be seen in a psychiatric hospital. Song et al. (Song et al., 2022) showed a decrease in incidence of MDD with increasing distance from CSJDM for outpatients, but not inpatients, supporting this view.

Our EHR-based Cox model identifies many of the same predictors found in registry-based studies performed in the Northern European countries of Denmark (Musliner & Ostergaard, 2018) and Finland (Baryshnikov et al., 2020). For example, as in Musliner & Ostergaard (Musliner & Ostergaard, 2018) and Baryshnikov et al., (Baryshnikov et al., 2020), we found gender and severity of the first MDD episode to be significantly related to conversion. The HR estimates of these consistently identified risk factors also very similar: males had lower rates of conversion to BD than did female (HR: 0.70-0.80); psychotic depression at the first episode increased rates of conversion (HR: 1.7-2.0). As in Musliner & Ostergaard (Musliner & Ostergaard, 2018), we further identify family history of mental illness, presence of psychosis at the first MDD, ER treatment or hospitalization at the first MDD, and a history of alcohol use/abuse before the first MDD to be significantly associated with the rate of conversion to BD. These similarities highlight the validity of using EHR for identifying risk factors of diagnostic changes.

Our study further builds on existing registry-based approaches quantifying the cumulative effect of the identified risk factors and their ability to predict conversion in a pre-defined time-frame, highlighting that EHR data can be used to predict disease trajectories. A recent study using insurance claims data (Nestsiarovich et al., 2021) observed similar predictive performance to our model; however, in their framework the prediction window was one year, and analyses were restricted to patients with complete follow-up and without a prior history of antipsychotic, antidepressant, lithium or mood-stabilizing drugs.

Predictive modeling from the complete EHR further allows the inclusion of additional features not commonly integrated with registry or claims data, such as symptoms and behaviors derived from clinical notes. To the best of our knowledge, we are the first to identify suicidal ideation and suicide attempts reported in clinical notes as risk factors for conversion to BD. These variables were identified as risk factors even when controlling for hospitalization at the index MDD visit. In addition, from available prescription data, we find the use of anti-psychotic medications and mood-stabilizing medications to increase the rate of conversion to BD, when controlling for diagnostic codes; conversely, in our data, anti-depressant us is protective factor.

Anti-psychotic use was also identified in Pradier (Pradier et al., 2021), a study that estimated risk factors for transition to BD within 90 days of the first prescription for an anti-depressant. Unlike our study, which relies on data from a psychiatric clinic and includes only patients treated by specialists, their work focused on general health care institutions. Indeed, the strongest risk factor observed in their study was being seen by a psychiatric provider (about 9% of their total sample), which increased the transition rate to BD compared to general care 3.5-fold.

Our finding that antidepressant use is associated with lower probability of conversion to BD appears unexpected, given the known risk of antidepressant-induced mania without concurrent mood stabilization therapy (Viktorin et al., 2014). In a predictive setting, the association between antidepressant use and conversion to BD has not been extensively studied and based on the few studies that exist, there is no clear consensus in the literature (Jo et al., 2022; Kim et al., 2020; Pacchiarotti et al., 2013). However, when evaluating this finding further, we observed that participants prescribed anti-depressants at the time of their first MDD episode are less severe: they are less likely to have been hospitalized at that first episode, less likely to have had a severe MDD diagnosis, less likely to have had a suicide attempt, and less likely to experience delusions. This confounding may account for the observed predictive effect, even when accounting for all other variables.

Another novelty of our study is our in-depth analyses of the role of prescriptions of mood stabilizers in conversion to BD. Our finding that the use of mood-stabilizing medication was predictive of conversion to BD could indicate that the attending physicians were cognizant of an increased risk of mania in these patients. Controlling for all the predictors used in our Cox model, the odds of being prescribed a mood stabilizer for patients with a family history of BD was 1.48x the odds for patients without a family history of BD, which could indicate that that physicians were using this information in developing their treatment plan. When we exclude the use of mood-stabilizing medications at the time of their first MDD as a predictor in our Cox regression model, we find that the model is still predictive of conversion to BD in the held-out data, with recall of 63% and AUC of 65%.

Evaluation of data from interim clinic visits, for patients who had multiple visits before conversion/censoring, indicated that HR were very similar for all predictors except for mood-stabilizer use. Patients who were not on mood stabilizers at their initial MDD visit, but subsequently were prescribed them, were identified as being at increased risk for conversion. Mood stabilizer use, however, is clearly well-known to physicians, who are likely already aware of the risk for conversion to BD for these patients. Other risk factors identified in our model had similar HR when using data from interim clinic visits as when using data from the first MDD episode, suggesting that accuracy of prediction of conversion is similar in the two approaches and conversion risk can be well estimated using data available at the time of the first MDD episode.

When testing our predictions in a temporally independent test set of 50 people who came for their second visit in 2022, we were able to correctly predict conversion by five years in all three converters who were observed to convert within five years.

## Limitations

A limitation of using EHR as opposed to registry-data is incomplete information: even in a setting such as here, when a caption area is well-defined, one can never know whether absence of recorded visits mean people left the region, stopped needing/using treatment, or passed away. Second, EHR have the issue of censoring; it is possible that patients convert to BD before the records started or after the follow-up time. Interestingly, we show that for every 10% increase in predicted probability of conversion, we observe 87 days shorter follow-up time in non-converters, thus indicating that that while patients did not convert during the follow-up time, they may still convert in the future, likely contributing to a modest AUC. Finally, while our EHR data are very detailed and complete, we are unable to link data recorded in the hospital with data from primary care providers.

## Conclusions

We showed that EHRs can be used to predict conversion from unipolar depression to bipolar disorder using data from a psychiatric hospital in Colombia. We replicate several risk factors of conversion to BD previously identified in patient registries and EHRs from upper income countries, and also identify novel such features: namely, suicidal ideation and suicide attempt at or before the index depressive episode. Using our multivariate model, we can identify patients at increased risk of conversion from MDD to BD. While our predictions are not yet at the level of clinically utility, we hypothesize that future work including expanded NLP libraries, genetic risk factors, and more complex temporal modelling will improve prediction.

## Data Availability

NA

## Acknowledgements

Research reported here was supported by R00MH116115 (to LMOL), R01MH123157 (to LMOL, CLJ, and NBF) and R01MH113078 (to CEB, CLJ, and NBF)

## Figure Legends

Supplementary Figure 1. Flow chart describing patient exclusion and formation of final analysis sample

Supplementary Figure 2. Kaplan-Meier survival curve of the entire data set (1,610 converters and 11,997 censored patients).

Supplementary Figure 3. Plot of scaled Schoenfeld residuals vs. time for use of mood stabilizing drugs (MS) in the Cox model applied to the training data.

Supplementary Figure 4. Distribution of the days of follow-up data for 4,082 patients in the test data, divided by quartile of the probability to convert to BD within five years of their initial MDD diagnosis, and whether or not the patient was observed to convert to BD or was censored. Q1=first quartile, Q2=second quartile, Q3=third quartile, Q4=fourth quartile.

Supplementary Figure 5. Recall-FPR plot (A) and Precision-Recall plot (B) estimated from application of results of the baseline Cox model in training data, applied to patients in the test data.

Supplementary Figure 6. Distribution of the probability to convert to BD within five years of the initial MDD diagnosis in 4,082 patients in the test data, using hazard ratios estimated in the training data, stratified by status (Censored/Convert) and mood stabilizer use at the time of the first MDD diagnosis.

Supplementary Figure 7. Hazard ratios and 95% confidence intervals from the baseline multivariate Cox model used on the training data, excluding individuals on mood stabilizers at the time of their first MDD diagnosis.

Supplementary Figure 8. Comparison of hazard ratios for predictors evaluated after the first MDD diagnosis in the baseline model and the time-dependent covariate model.

Supplementary Figure 9. Distribution of the probability to convert to BD within five years of the initial MDD diagnosis in 50 patients with a second visit to the clinic in 2022, using hazard ratios estimated in the training data.

