## Supplementary Figures Table for "Predicting diagnostic conversion from major depressive disorder to bipolar disorder: an EHR based study from Colombia"

Supplementary Figure 1

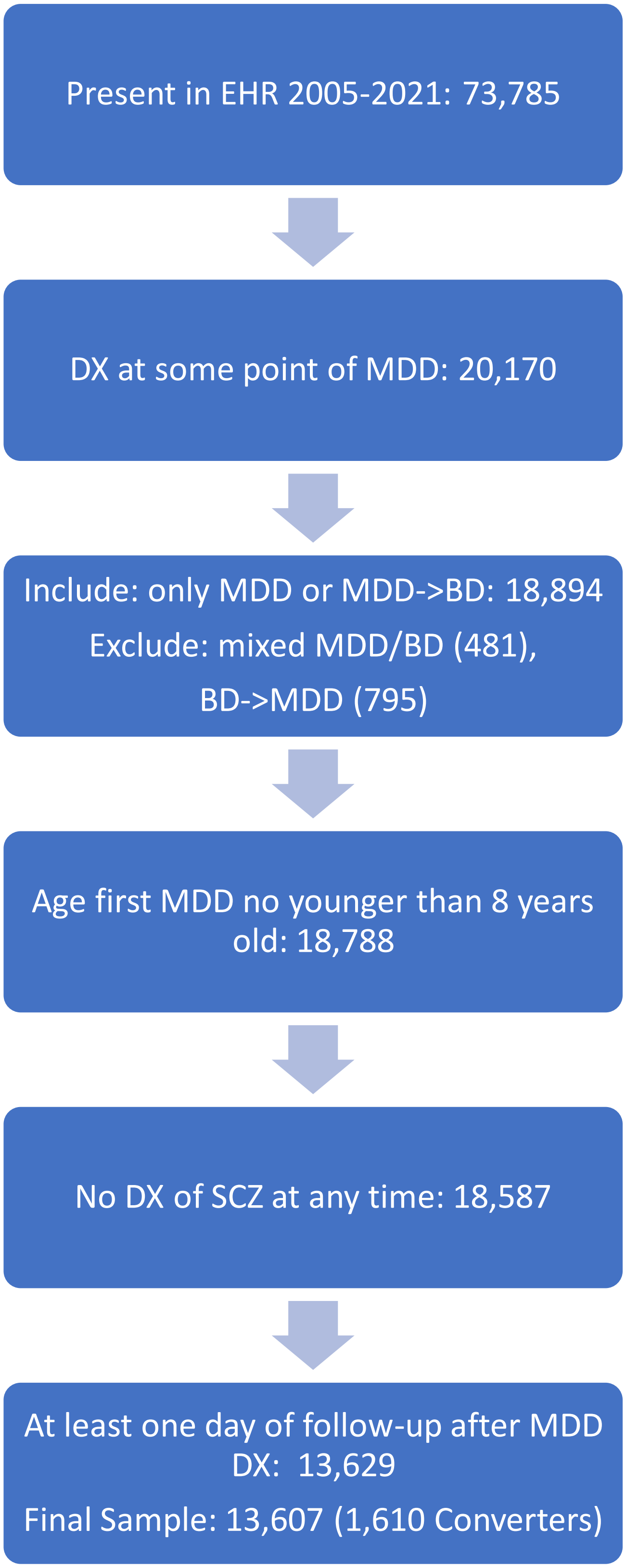

### Supplementary Figure 2

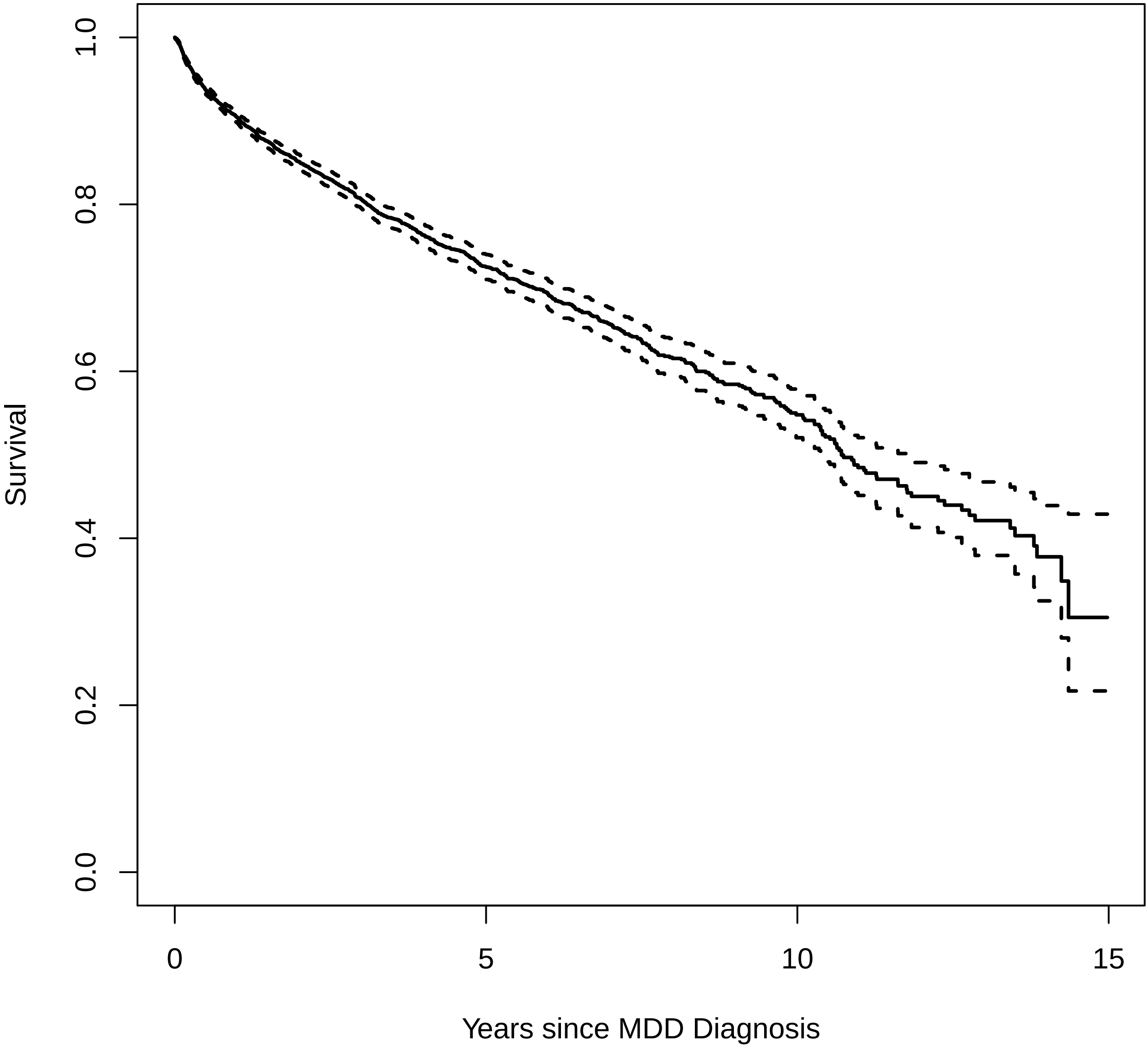

### Supplementary Figure 3

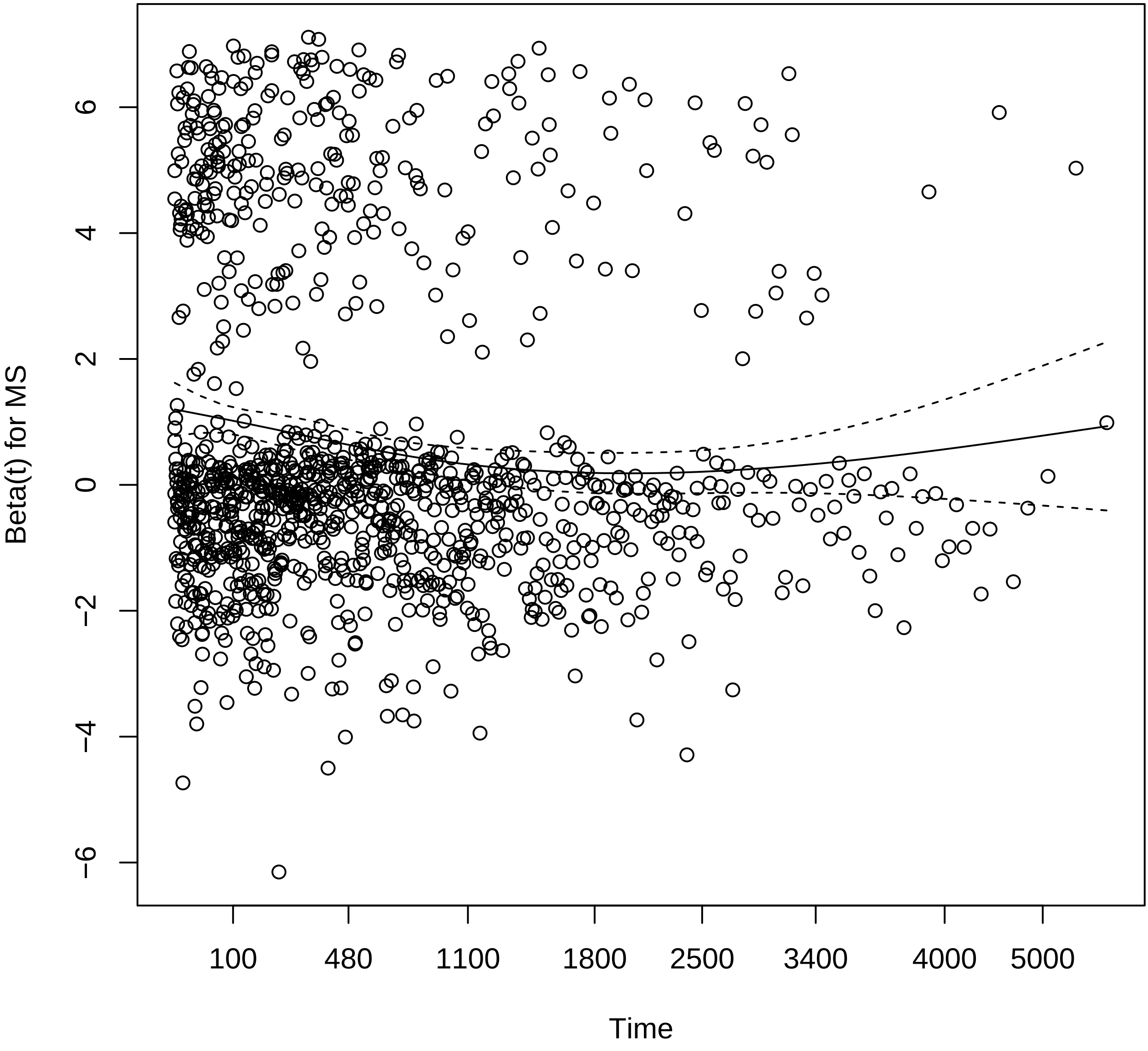

### Supplementary Figure 4

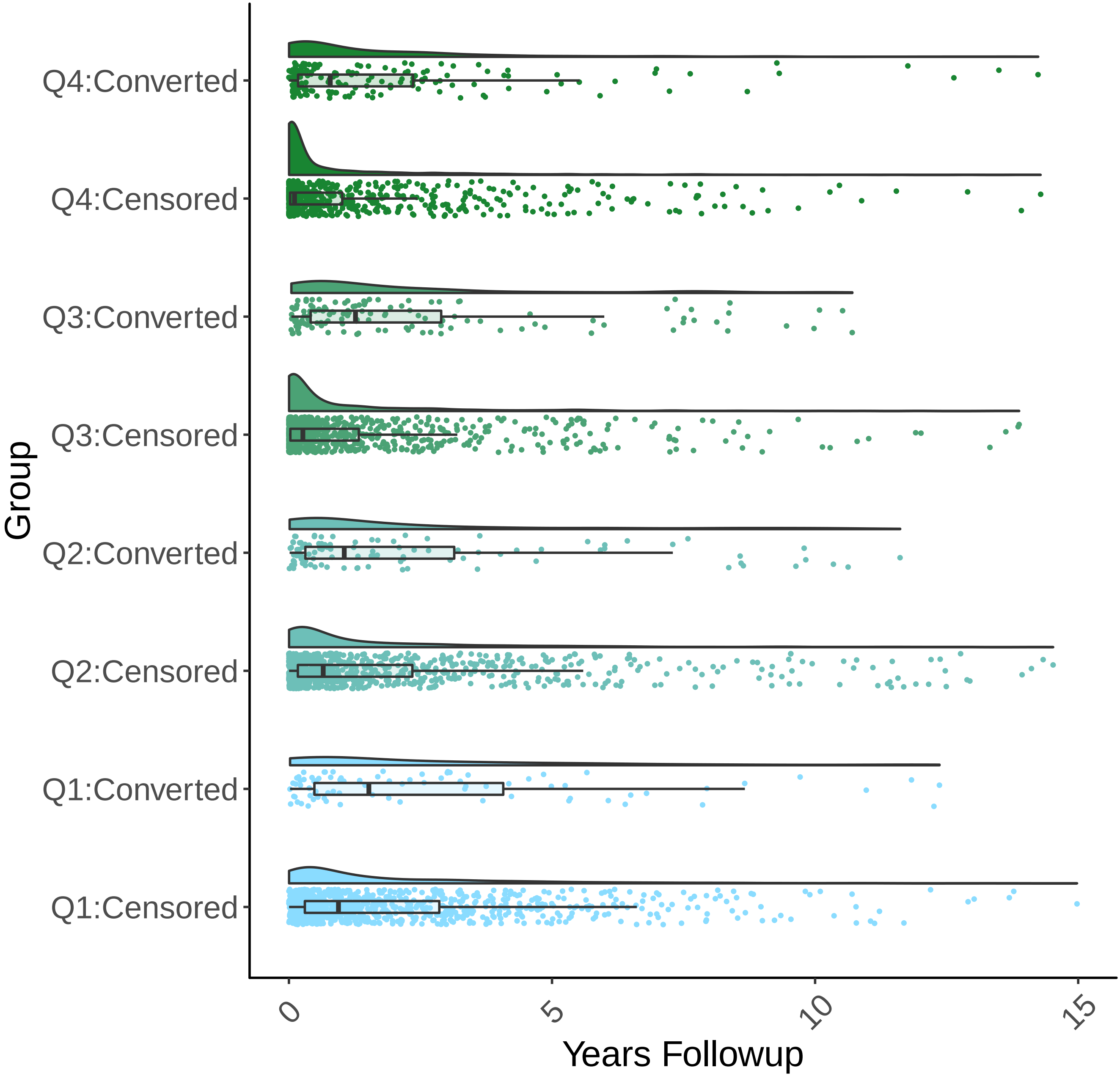

Supplementary Figure 5A

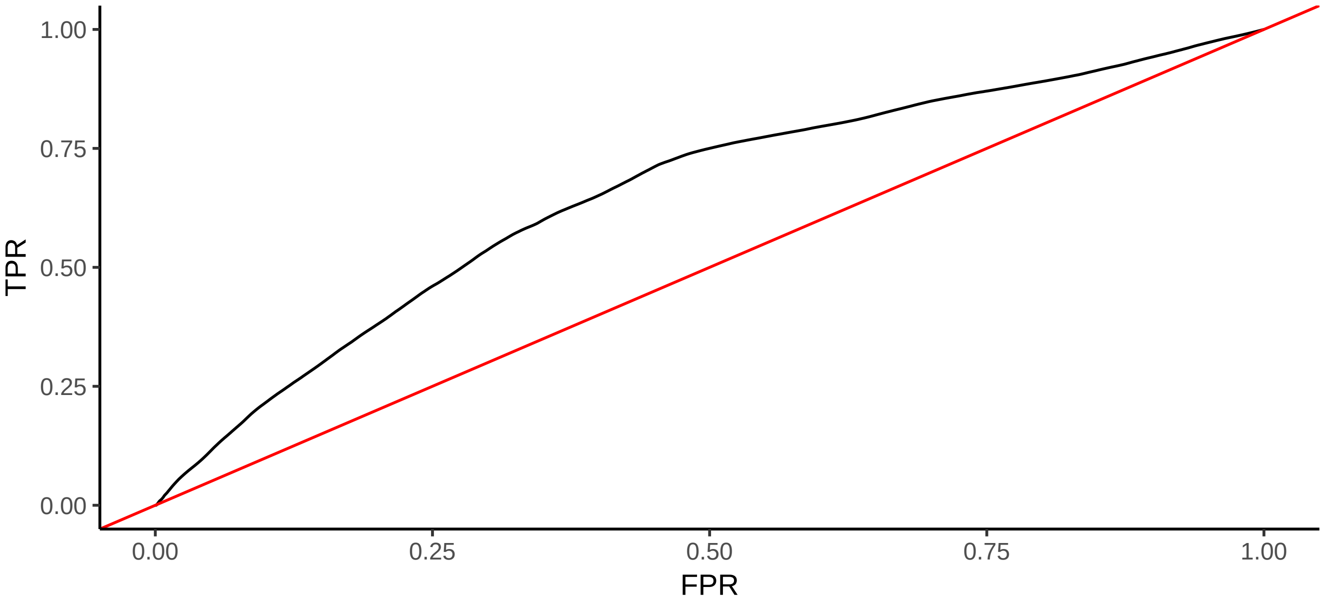


Supplementary Figure 5B

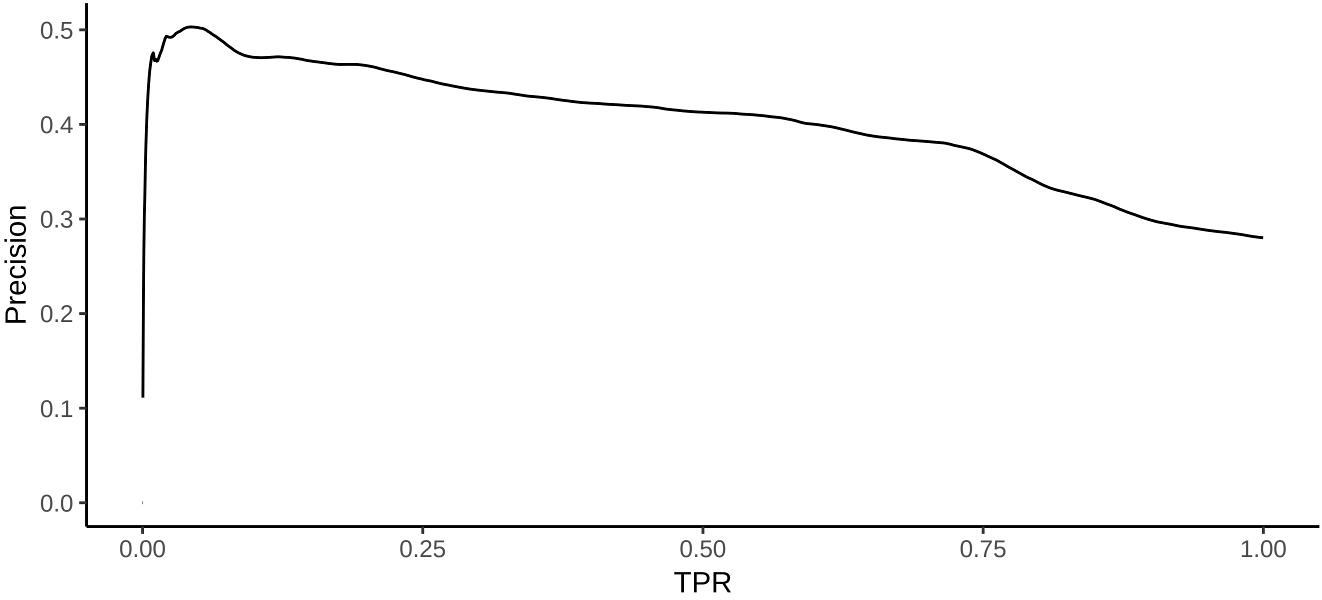

### Supplementary Figure 6

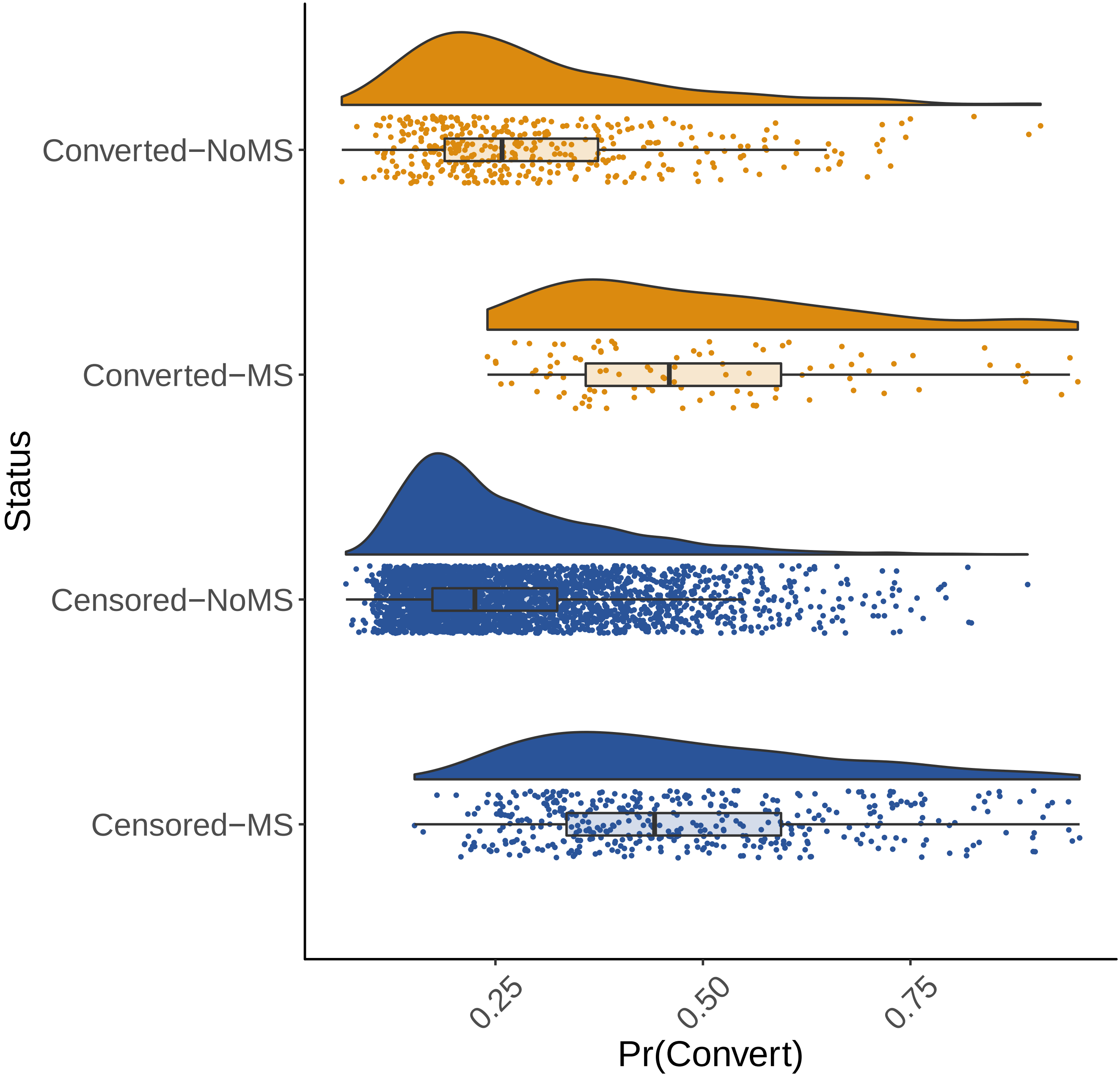

### Supplementary Figure 7

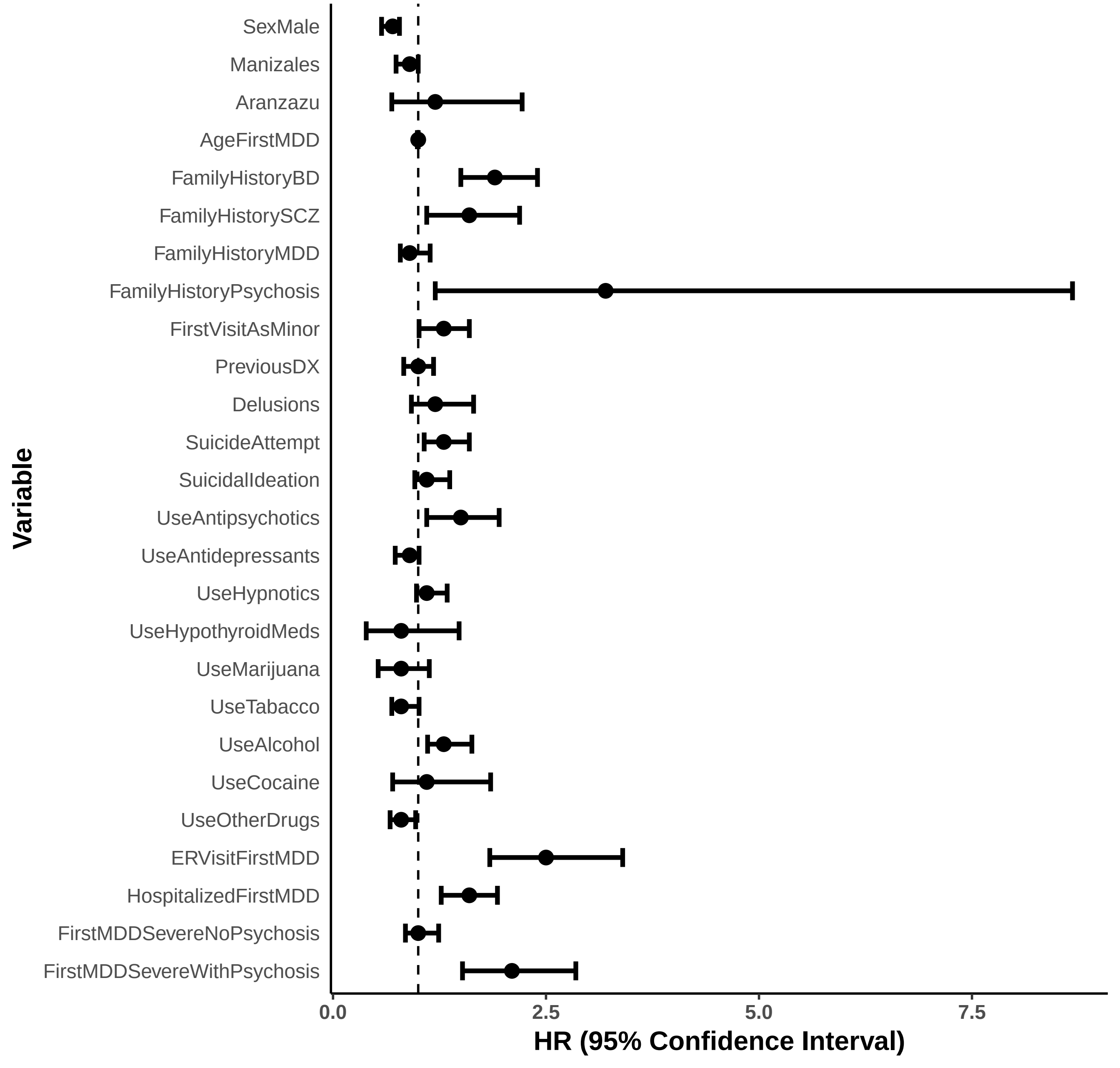

### Supplementary Figure 8A

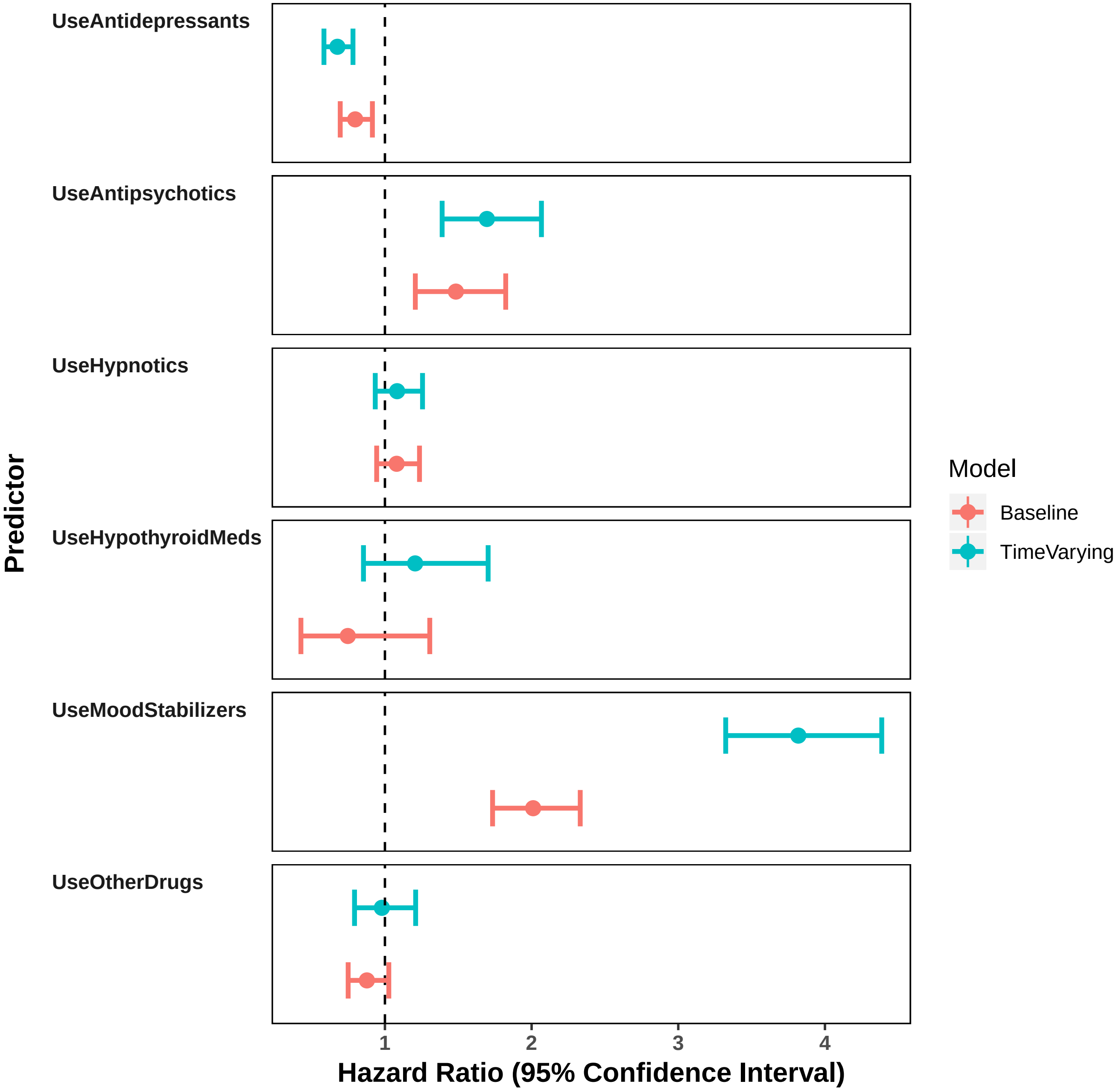

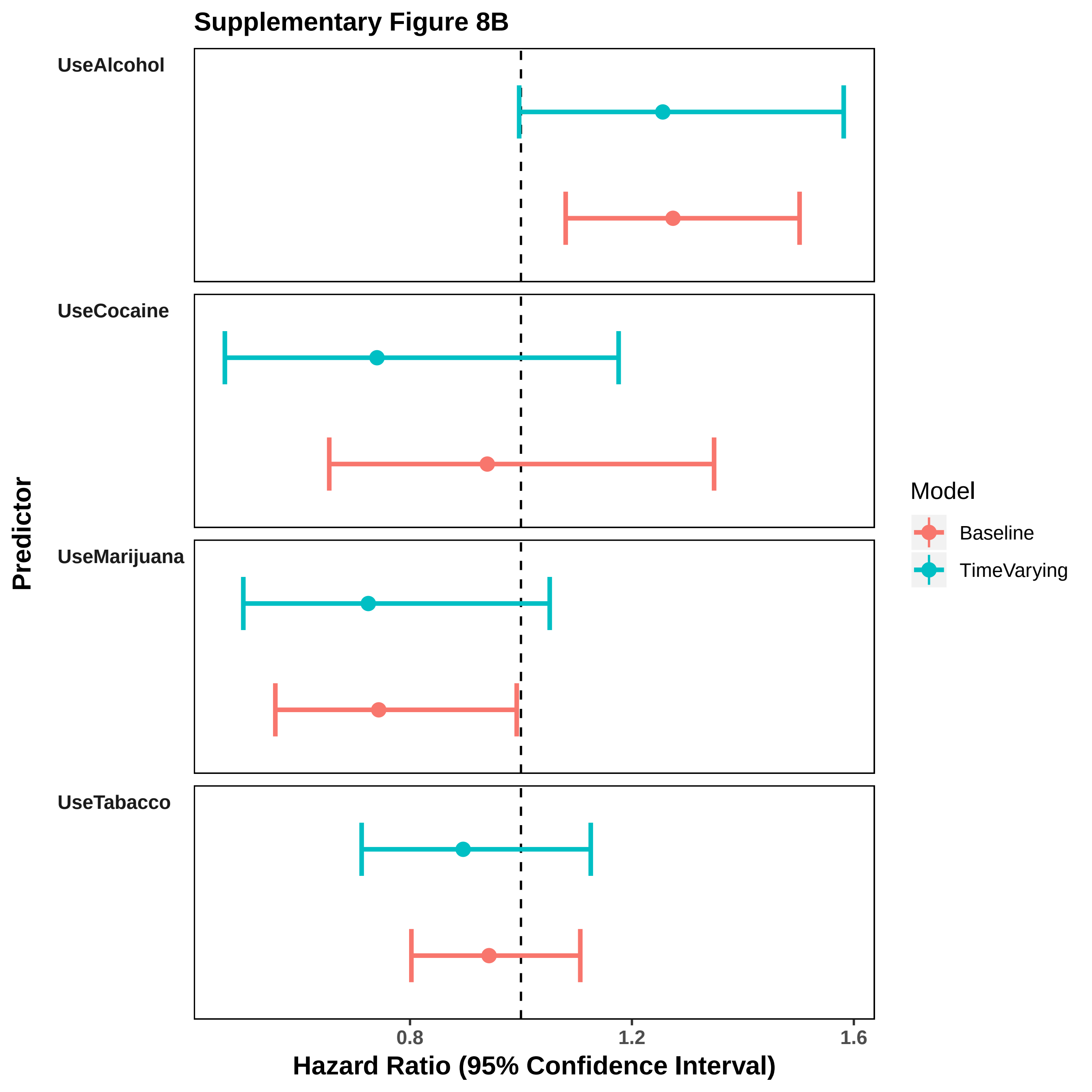
Supplementary Figure 8B
UseAlcohol
UseCocaine
UseMarijuana
UseTabacco
Predictor
Model
Baseline
TimeVarying
0.8
1.2
1.6
Hazard Ratio (95% Confidence Interval)

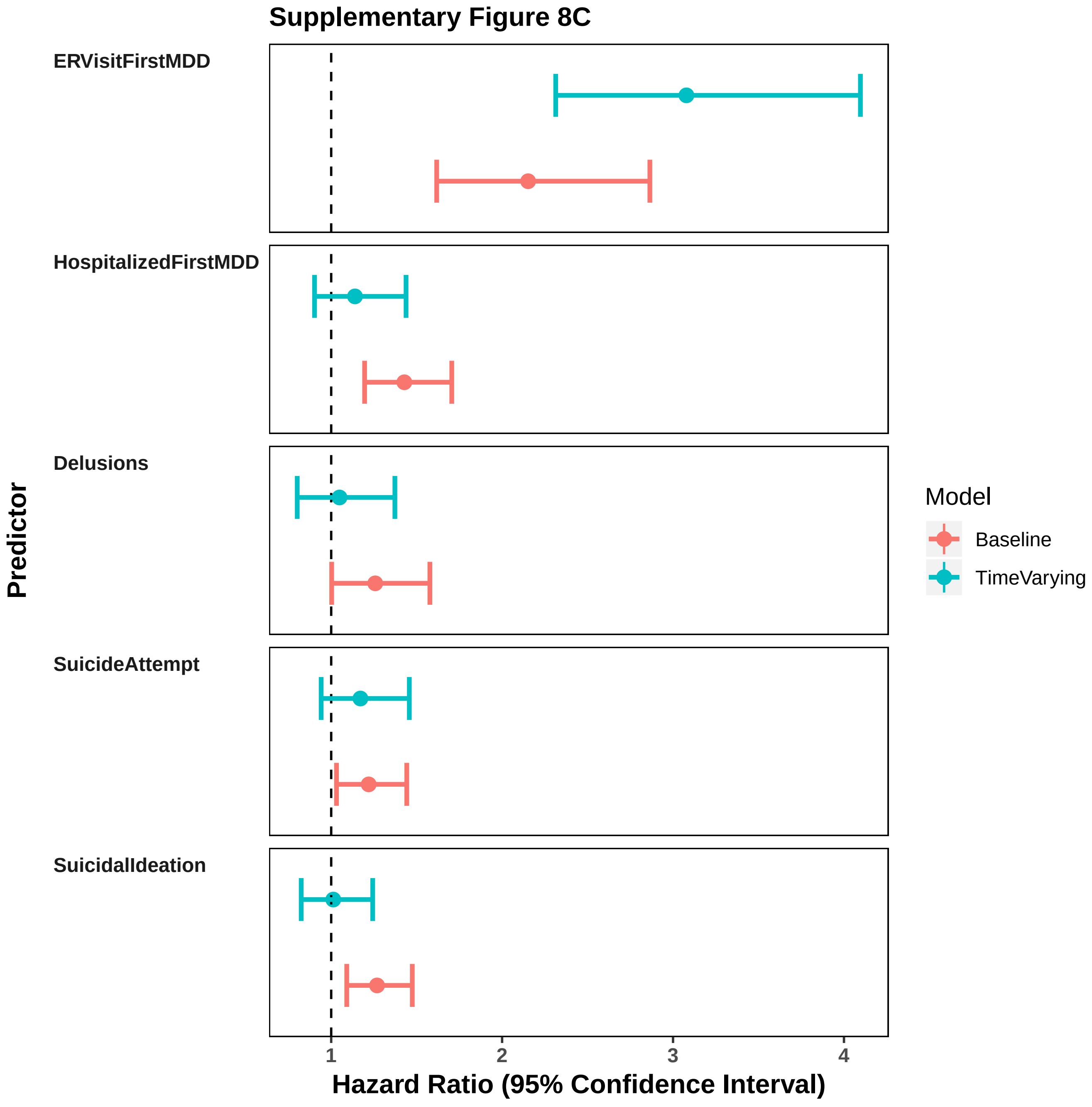
Supplementary Figure 8C
ERVisitFirstMDD
HospitalizedFirstMDD
Delusions
SuicideAttempt
SuicidalIdeation
Predictor
1
2
3
4
Hazard Ratio (95% Confidence Interval)
Model
Baseline
TimeVarying

### Supplementary Figure 9

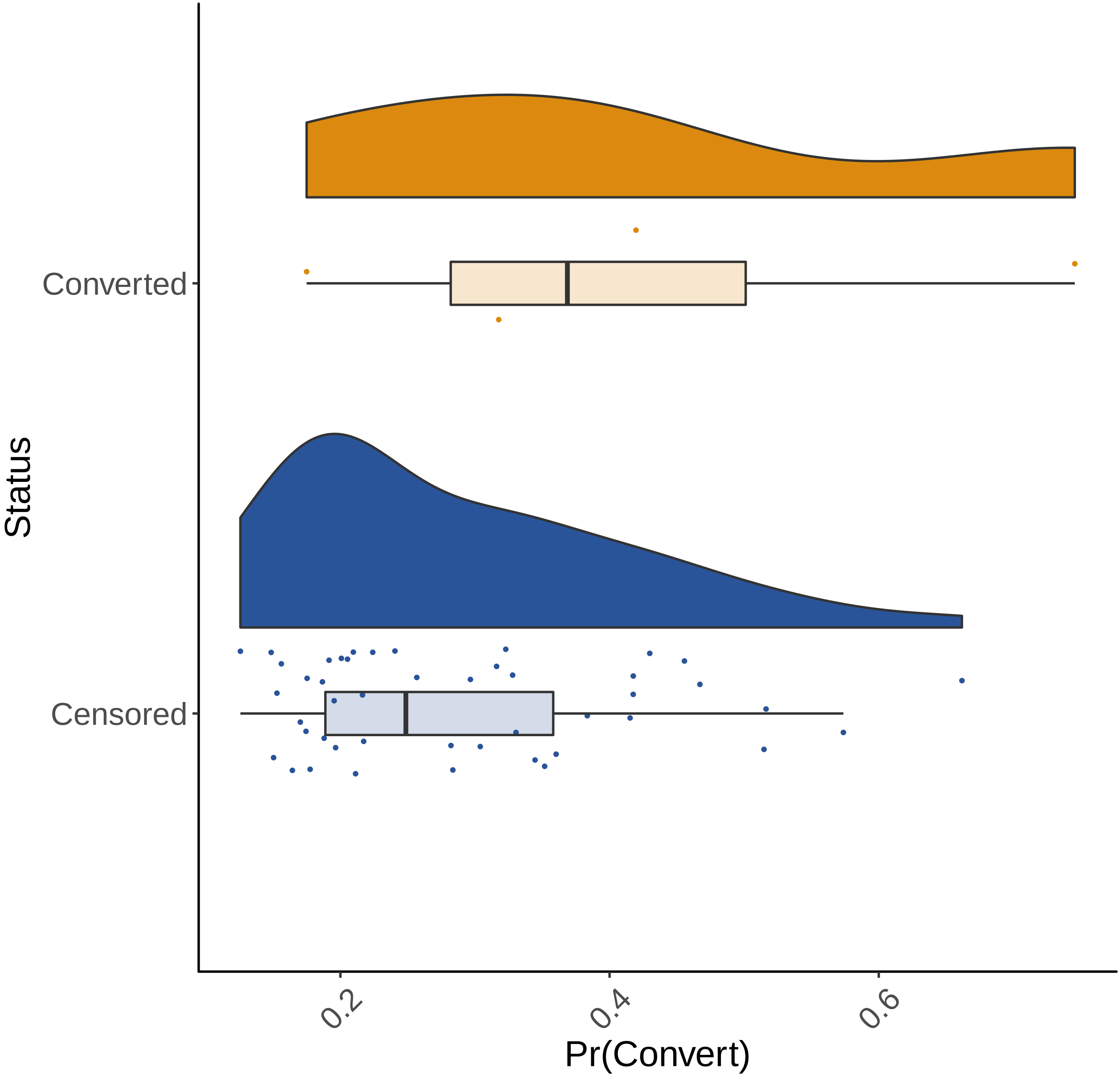

Supplementary Table 1. Hazard Ratio (HR), lower (LL.CI) and upper (UL.CI) 95% confidence intervals and p-value (P) for the null hypothesis that the HR=1. Results are from the multivariate model applied to the training data

| **Variable** | **HR** | **LL.CI** | **UL.CI** | **P** |
|---|---|---|---|---|
| SexMale | 0.7 | 0.61 | 0.79 | 8.05E-08 |
| Manizales | 0.8 | 0.73 | 0.94 | 3.60E-03 |
| Aranzazu | 1.4 | 0.88 | 2.11 | 1.62E-01 |
| AgeFirstMDD | 1 | 0.99 | 1 | 2.36E-01 |
| FamilyHistoryBD | 1.8 | 1.47 | 2.23 | 2.00E-08 |
| FamilyHistorySCZ | 1.5 | 1.08 | 1.98 | 1.45E-02 |
| FamilyHistoryMDD | 0.9 | 0.81 | 1.11 | 5.01E-01 |
| FamilyHistoryPsychosis | 1.7 | 0.72 | 3.93 | 2.33E-01 |
| FirstVisitAsMinor | 1.3 | 1.03 | 1.52 | 2.41E-02 |
| PreviousDX | 0.9 | 0.75 | 1.02 | 8.30E-02 |
| Delusions | 1.3 | 1 | 1.58 | 4.71E-02 |
| SuicideAttempt | 1.2 | 1.03 | 1.44 | 2.05E-02 |
| SuicidalIdeation | 1.3 | 1.09 | 1.47 | 1.98E-03 |
| UseAntipsychotics | 1.5 | 1.21 | 1.82 | 1.76E-04 |
| UseAntidepressants | 0.8 | 0.7 | 0.91 | 1.21E-03 |
| UseMoodStabilizers | 2 | 1.73 | 2.33 | 2.31E-20 |
| UseHypnotics | 1.1 | 0.94 | 1.24 | 2.64E-01 |
| UseHypothyroidMeds | 0.7 | 0.43 | 1.31 | 3.06E-01 |
| UseMarijuana | 0.7 | 0.56 | 0.99 | 4.42E-02 |
| UseTabacco | 0.9 | 0.8 | 1.11 | 4.70E-01 |
| UseAlcohol | 1.3 | 1.08 | 1.5 | 3.95E-03 |
| UseCocaine | 0.9 | 0.65 | 1.35 | 7.34E-01 |
| UseOtherDrugs | 0.9 | 0.75 | 1.03 | 1.02E-01 |
| ERVisitFirstMDD | 2.2 | 1.62 | 2.86 | 1.47E-07 |
| HospitalizedFirstMDD | 1.4 | 1.2 | 1.71 | 8.49E-05 |
| FirstMDDSevereNoPsychosis | 1 | 0.89 | 1.22 | 6.23E-01 |
| FirstMDDSevereWithPsychosis | 1.9 | 1.46 | 2.41 | 7.08E-07 |

Supplementary Table 2. Associations of covariates to antidepressant use

| **Variable** | **logOR** | **logOR_SE** | **P-value** |
|---|---|---|---|
| SexMale | 0.032 | 0.049 | 5.10E-01 |
| PreviousDX | 1.153 | 0.072 | 5.08E-58 |
| FirstVisitAsMinor | 0.090 | 0.056 | 1.10E-01 |
| ERVisitFirstMDD | 0.361 | 0.131 | 5.92E-03 |
| HospitalizedFirstMDD | -0.834 | 0.047 | 3.36E-71 |
| FirstMDDSevereWithPsychosis | -1.136 | 0.103 | 2.55E-28 |
| FirstMDDSevereNoPsychosis | -0.384 | 0.049 | 5.77E-15 |
| UseAntipsychotics | -0.046 | 0.096 | 6.29E-01 |
| UseMoodStabilizers | -0.082 | 0.068 | 2.31E-01 |
| SuicideAttempt | -0.271 | 0.056 | 1.64E-06 |
| SuicidalIdeation | 0.061 | 0.052 | 2.44E-01 |
| Delusions | -0.366 | 0.095 | 1.10E-04 |
| FamilyHistoryBD | -0.137 | 0.114 | 2.31E-01 |
| FamilyHistoryMDD | 0.297 | 0.072 | 3.31E-05 |
